# Astrocyte biomarker signatures of amyloid-β and tau pathologies in Alzheimer’s disease

**DOI:** 10.1101/2022.01.25.22269841

**Authors:** João Pedro Ferrari-Souza, Pâmela C. Lukasewicz Ferreira, Bruna Bellaver, Cécile Tissot, Yi-Ting Wang, Douglas T. Leffa, Wagner S. Brum, Andréa L. Benedet, Nicholas J. Ashton, Marco Antônio De Bastiani, Andréia Rocha, Joseph Therriault, Firoza Z. Lussier, Mira Chamoun, Stijn Servaes, Gleb Bezgin, Min Su Kang, Jenna Stevenson, Nesrine Rahmouni, Vanessa Pallen, Nina Margherita Poltronetti, William E. Klunk, Dana L. Tudorascu, Annie Cohen, Victor L. Villemagne, Serge Gauthier, Kaj Blennow, Henrik Zetterberg, Diogo O. Souza, Thomas K. Karikari, Eduardo R. Zimmer, Pedro Rosa-Neto, Tharick A. Pascoal

## Abstract

Astrocytes can adopt multiple molecular phenotypes in the brain of Alzheimer’s disease (AD) patients. Here, we studied the associations of cerebrospinal fluid (CSF) glial fibrillary acidic protein (GFAP) and chitinase-3-like protein 1 (YKL-40) levels with brain amyloid-β (Aβ) and tau pathologies. We assessed 121 individuals across the aging and AD clinical spectrum with positron emission tomography (PET) brain imaging for Aβ ([^18^F]AZD4694) and tau ([^18^F]MK6240), as well as CSF GFAP and YKL-40 measures. We observed that higher CSF GFAP levels were associated with elevated Aβ-PET but not tau-PET load. By contrast, higher CSF YKL-40 levels were associated with elevated tau-PET but not Aβ-PET burden. Structural equation modeling revealed that CSF GFAP and YKL-40 mediate the effects of Aβ and tau, respectively, on hippocampal atrophy, ultimately leading to cognitive impairment. Our results suggest the existence of distinct astrocyte biomarker signatures in response to brain Aβ and tau accumulation, which may contribute to our understanding of the complex link between reactive astrogliosis heterogeneity and AD progression.

## Introduction

Reactive astrocytes play an important role in Alzheimer’s disease (AD) pathophysiology^1-5^. Post-mortem studies suggest that both amyloid-β (Aβ) and tau pathologies are associated with astrocyte reactivity^1,6^. Far from displaying a homogenous response, transcriptomics analyses demonstrated that reactive astrocytes can acquire multiple molecular phenotypes in the AD brain^7^. The context-specific aspects of astrocyte reactivity^8^ raise the possibility that astrocytes respond differently to AD-related brain processes. In fact, experimental evidence indicates the presence of distinct but overlapping molecular astrocyte signatures in response to Aβ and tau pathologies^9^. However, knowledge about Aβ- and tau-specific contributions to reactive astrocyte biomarkers in patients with AD is still limited.

Reactive astrocytes overexpress specific proteins that can be released into the extracellular compartment, being measured in the cerebrospinal fluid (CSF) of living individuals^8,10^. CSF levels of glial fibrillary acidic protein (GFAP) and chitinase-3-like protein 1 (YKL-40), biomarkers of astrocyte reactivity^8,10^, are consistently elevated in the dementia phase of AD^11^, and in some other brain disorders such as multiple sclerosis^12,13^. Although GFAP and YKL-40 fluid concentrations have already been shown to correlate with AD pathophysiology^14-19^, no previous study has investigated the existence of Aβ-and tau-related astrocyte responses in the human brain. Identifying astrocyte biomarker signatures related to AD proteinopathies has the potential to provide insights into the role of astrocytes in disease progression, allow disease staging, and can lead to the development of drugs targeting distinct reactive astrocyte phenotypes.

In a cohort of individuals across the aging and AD clinical spectrum, we tested whether CSF GFAP and YKL-40 are distinctly associated with Aβ and tau pathologies. We also investigated whether the reactive astrocyte biomarkers mediate the effects of AD hallmark pathologies on neurodegeneration and cognitive impairment. Lastly, we tested whether CSF GFAP and YKL-40 are related to CSF inflammatory markers.

## Results

We studied 75 cognitively unimpaired (CU) and 46 cognitively impaired [CI; 29 with mild cognitive impairment (MCI) and 17 with AD dementia] individuals from 52 to 85 years of age. Demographic information of the population can be found in **Table 1**. No statistically significant difference was observed between CU and CI groups regarding age, sex, and years of education. CI subjects had lower Mini-Mental State Examination (MMSE) scores, higher neocortical Aβ-PET standardized uptake value ratio (SUVR), temporal meta-region of interest (ROI) tau-PET SUVR, and lower hippocampal volume than CU subjects. Additionally, individuals in the CI group were more likely to be apolipoprotein E ε4 (*APOE* ε4) carriers compared to individuals in the CU group. Correlations between reactive astrocyte biomarkers are presented in **Supplementary Fig. 1**.

**Table 1.** Demographics and key characteristics of participants by cognitive status.

|  | CU | CI | *P*-value |
| --- | --- | --- | --- |
| No. | 75 | 46 | - |
| Age, years | 70.9 (5.8) | 68.9 (7.6) | 0.136 |
| Male, No. (%) | 28 (37.3) | 25 (54.3) | 0.101 |
| Education, years | 14.7 (3.4) | 15.3 (3.1) | 0.359 |
| *APOE* ε4 carriers, No. (%) | 22 (29.3) | 26 (56.5) | 0.006 |
| MMSE score | 29.1 (1.0) | 25.2 (5.4) | < 0.001 |
| Neocortical Aβ-PET SUVR | 1.51 (0.4) | 2.20 (0.6) | < 0.001 |
| Temporal meta-ROI tau-PET SUVR | 0.86 (0.1) | 1.59 (0.8) | < 0.001 |
| Hippocampal volume, cm<sup>3</sup> | 3.53 (0.4) | 3.16 (0.5) | < 0.001 |

Continuous variables are presented as mean (SD). Aβ = amyloid-β ; *APOE* ε4 = Apolipoprotein E ε4; CI = cognitively impaired; CSF = cerebrospinal fluid; CU = cognitively unimpaired; GFAP = glial fibrillary acidic protein; MMSE = Mini-Mental State Examination; ROI = region of interest; SD = standard deviation; SUVR = standardized uptake value ratio; YKL-40 = chitinase-3-like protein 1.

### Higher GFAP levels are associated with Aβ positivity and YKL-40 levels with tau positivity

We assessed the levels of reactive astrocyte biomarkers across groups defined by Aβ-PET (A) and tau-PET (T) status. CSF GFAP levels were significantly higher in A+T- and A+T+ groups compared with the A-T-group (A+T-*vs*. A-T-: *P* = 0.048; A+T+ *vs*. A-T-: *P* < 0.001; **Fig. 1A**). Furthermore, no statistically significant difference was observed between A+T- and A+T+ groups (*P* = 0.196; **Fig. 1A**). In relation to CSF GFAP, similar findings were observed for plasma GFAP levels across groups (**Supplementary Fig. 2A**). CSF YKL-40 levels were higher in the A+T+ group as compared with A-T-(*P* = 0.006) and A+T-(*P* = 0.004) groups (**Fig. 1B**). Moreover, no significant difference was observed when comparing A-T- and A+T-groups (*P* = 0.868; **Fig. 1B**).

**Figure 1.**
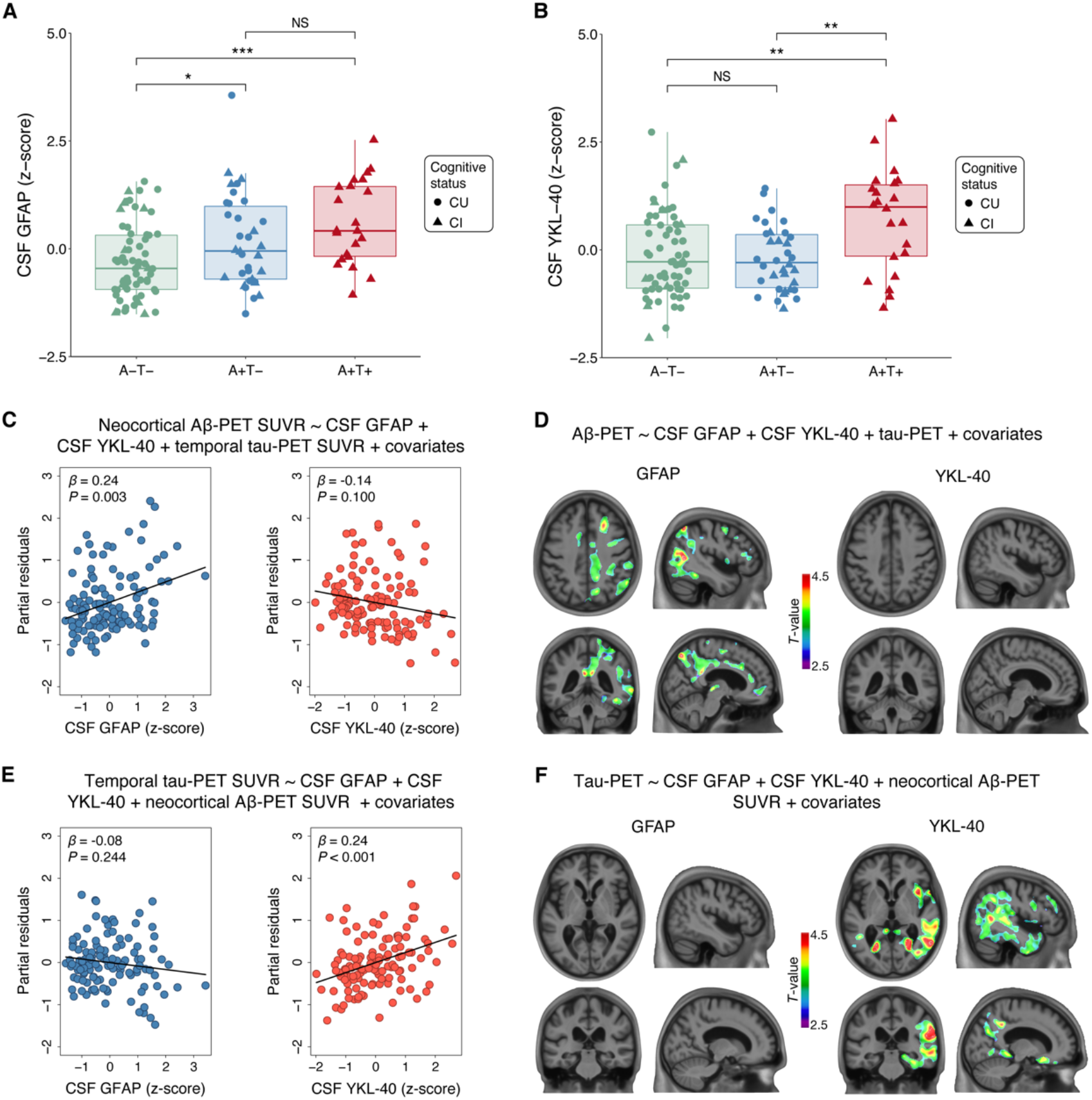
GFAP associates with Aβ and YKL-40 with tau accumulation. The panels show box-and-whisker plots of (**A**) CSF GFAP and (**B**) CSF YKL-40 levels adjusted for age-, sex-, and *APOE* ε4 status across AT groups. The horizontal line in each box represents the median; box ends represent the 25th and 75th percentiles. Groups were compared using analyses of variance with Tukey’s multiple comparison test (\**P*<0.05, \*\**P*<0.01, \*\*\**P*<0.001). (**C**) Partial residual plots of ROI-based linear regressions testing the associations of neocortical Aβ-PET SUVR with CSF GFAP and YKL-40 levels adjusting for temporal meta-ROI tau-PET SUVR. (**D**) T-statistical parametric maps show the result of voxel-wise linear regression testing the regional association of Aβ-PET SUVR with CSF GFAP and YKL-40 levels adjusting for tau-PET SUVR. (**E**) Partial residual plots of ROI-based linear regressions testing the associations of temporal meta-ROI tau-PET SUVR with CSF GFAP and YKL-40 levels adjusting for neocortical Aβ-PET SUVR. (**F**) T-statistical parametric maps show the result of voxel-wise linear regression testing the regional association of tau-PET SUVR with CSF GFAP and YKL-40 levels adjusting for neocortical Aβ-PET SUVR. Voxel-wise linear regressions were RFT-corrected for multiple comparisons at a voxel threshold of *P* < 0.001. Age, sex, cognitive status, and *APOE* ε4 status were used as covariates for adjustment in all ROI-and voxel-based linear regressions. NS = not significant.

### GFAP but not YKL-40 is associated with Aβ-PET burden

We investigated the associations of Aβ-PET burden with CSF GFAP and YKL-40 levels adjusting for tau-PET SUVR, age, sex, cognitive status, and *APOE* ε4 status. ROI-based linear regression model revealed that higher CSF GFAP but not CSF YKL-40 levels were associated with higher neocortical Aβ-PET SUVR (CSF GFAP: β = 0.24, *P* = 0.003; CSF YKL-40: β = -0.14, *P* = 0.100; **Fig. 1C** and Model A on **Supplementary Table 1**). Voxel-wise analysis showed that CSF GFAP levels were positively associated with Aβ-PET load in Aβ-related brain regions (*e*.*g*., precuneus, cingulate, orbitofrontal, and lateral temporal; **Fig. 1D**). No association was detected between CSF YKL-40 concentrations and Aβ-PET SUVR (**Fig. 1D**). We conducted sensitivity analysis using plasma GFAP instead of CSF GFAP. We observed that plasma GFAP but not CSF YKL-40 levels were positively associated with Aβ-PET burden (plasma GFAP: β = 0.35, *P* < 0.001; CSF YKL-40: β = 0.05, *P* = 0.488; **Supplementary Fig. 2B**), reinforcing the aforementioned results.

### YKL-40 but not GFAP is associated with tau-PET burden

We further tested the associations of tau-PET uptake with CSF GFAP and YKL-40 levels adjusting for Aβ-PET SUVR, age, sex, cognitive status, and *APOE* ε4 status. ROI-based regressions demonstrated that higher CSF YKL-40 but not CSF GFAP levels were associated with higher temporal meta-ROI tau-PET SUVR (CSF GFAP: β = -0.08, *P* = 0.244; CSF YKL-40: β = 0.24, *P* < 0.001; **Fig. 1E** and Model B on **Supplementary Table 1**). Voxel-wise linear regression analysis confirmed that CSF YKL-40 levels were positively associated with tau-PET uptake in early and late Braak regions (**Fig. 1F**). No association was observed between CSF GFAP levels and tau-PET uptake (**Fig. 1F**). In sensitivity analysis using plasma GFAP instead of CSF GFAP, we found that higher CSF YKL-40 but not plasma GFAP levels were associated with higher tau-PET burden (plasma GFAP: β = 0.07, *P* = 0.342; CSF YKL-40: β = 0.16, *P* = 0.012; **Supplementary Fig. 2C**), which supports the aforementioned results.

### GFAP and YKL-40 mediate hippocampal atrophy and cognitive impairment

We tested whether CSF GFAP mediates the association of AD pathophysiology with hippocampal atrophy and cognitive impairment using structural equation modeling. The model demonstrated that CSF GFAP levels partially mediate the effect of higher Aβ-PET on lower hippocampal volume. We observed an indirect effect of Aβ pathology on cognition through increasing CSF GFAP levels and decreasing hippocampal volume (**Fig. 2A**). The aforementioned structural equation model fitted the data well [comparative fit index (CFI) = 1.00; root mean squared error of approximation (RMSEA) = 0.00, 90% confidence interval: 0.00-0.00; standardized root mean square residual (SRMR) = 0.00]. See **Supplementary Table 2** for complete model coefficients and associated statistics.

**Figure 2.**
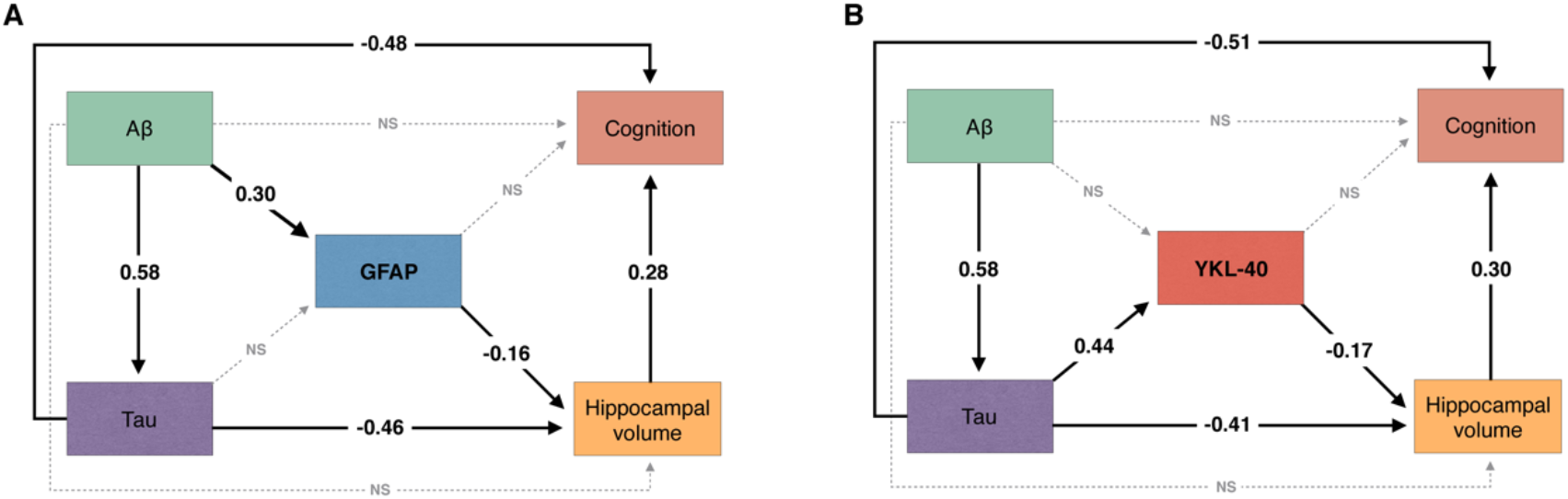
Reactive astrocyte biomarkers mediate the effect of AD pathophysiology on downstream neurodegeneration and cognitive impairment. The figure shows the standardized structural equation model estimates of the associations between (**A**) CSF GFAP and (**B**) CSF YKL-40 levels, hippocampal volume, and cognition. Solid black lines represent significant associations, whereas dashed gray lines represent non-significant effects. The fact that the presented estimates are standardized allows direct comparison between effects. Aβ pathology was measured with neocortical Aβ-PET SUVR and tau pathology using temporal meta-ROI tau-PET SUVR. Cognition was indexed using MMSE score. All associations were further adjusted for age, *APOE* ε4 status, and years of education. NS = not significant.

Furthermore, we tested whether CSF YKL-40 mediates the association of AD pathophysiology with hippocampal atrophy and cognitive impairment using structural equation modeling. The model revealed that higher tau-PET uptake was associated with lower hippocampal volume directly as well as indirectly through increasing CSF YKL-40 levels. The model suggests that tau effects on cognitive impairment were partially mediated through increasing CSF YKL-40 levels and reducing hippocampal volume (**Fig. 2B**). The aforementioned structural equation model yielded a robust fit (CFI = 1.00; RMSEA = 0.00, 90% confidence interval: 0.00-0.00; SRMR = 0.00). See **Supplementary Table 3** for model coefficients and associated statistics.

### Reactive-astrocyte biomarkers are associated with brain inflammation

In a subgroup of 62 participants (36 CU and 26 CI), we assessed the associations of CSF GFAP and YKL-40 with several inflammation-related proteins. Demographics for the subgroup of participants can be found in **Supplementary Table 4**. We observed that both CSF GFAP and YKL-40 were positively correlated with CSF inflammatory markers, including eukaryotic translation initiation factor 4E-binding protein 1 (4E-BP1), macrophage colony-stimulating factor 1 (CSF-1), fractalkine (CX3CL1), FMS-related tyrosine kinase 3 ligand (Flt3L), hepatocyte growth factor (HGF), interleukin-10 receptor subunit beta (IL-10RB), leukemia inhibitory factor receptor (LIF-R), matrix metalloproteinase-10 (MMP-10), stem cell factor (SCF), STAM-binding protein (STAMBP), transforming growth factor alpha (TGF-alpha), TNF-related apoptosis-inducing ligand (TRAIL), tumor necrosis factor ligand superfamily member 12 (TWEAK), and urokinase-type plasminogen activator (uPA) (**Fig. 3A, B**). Additionally, CSF YKL-40 but not CSF GFAP was also positively correlated with CD40L receptor (CD40), T-cell surface glycoprotein CD5 (CD5), C-X-C motif chemokine 1 (CXCL1), interleukin-12 subunit beta (IL-12B), interleukin-8 (IL-8), SIR2-like protein 2 (SIRT2), and vascular endothelial growth factor A (VEGF-A; **Fig. 3B**). No significant negative associations of CSF GFAP and YKL-40 with inflammatory markers were found, supporting the positive association of reactive astrocyte markers with inflammatory proteins in the CSF. A protein-protein interaction network confirmed that GFAP and YKL-40 proteins were inter-connected with the majority (19 out of 21) of the inflammation-related proteins (**Fig. 3C**).

**Figure 3.**
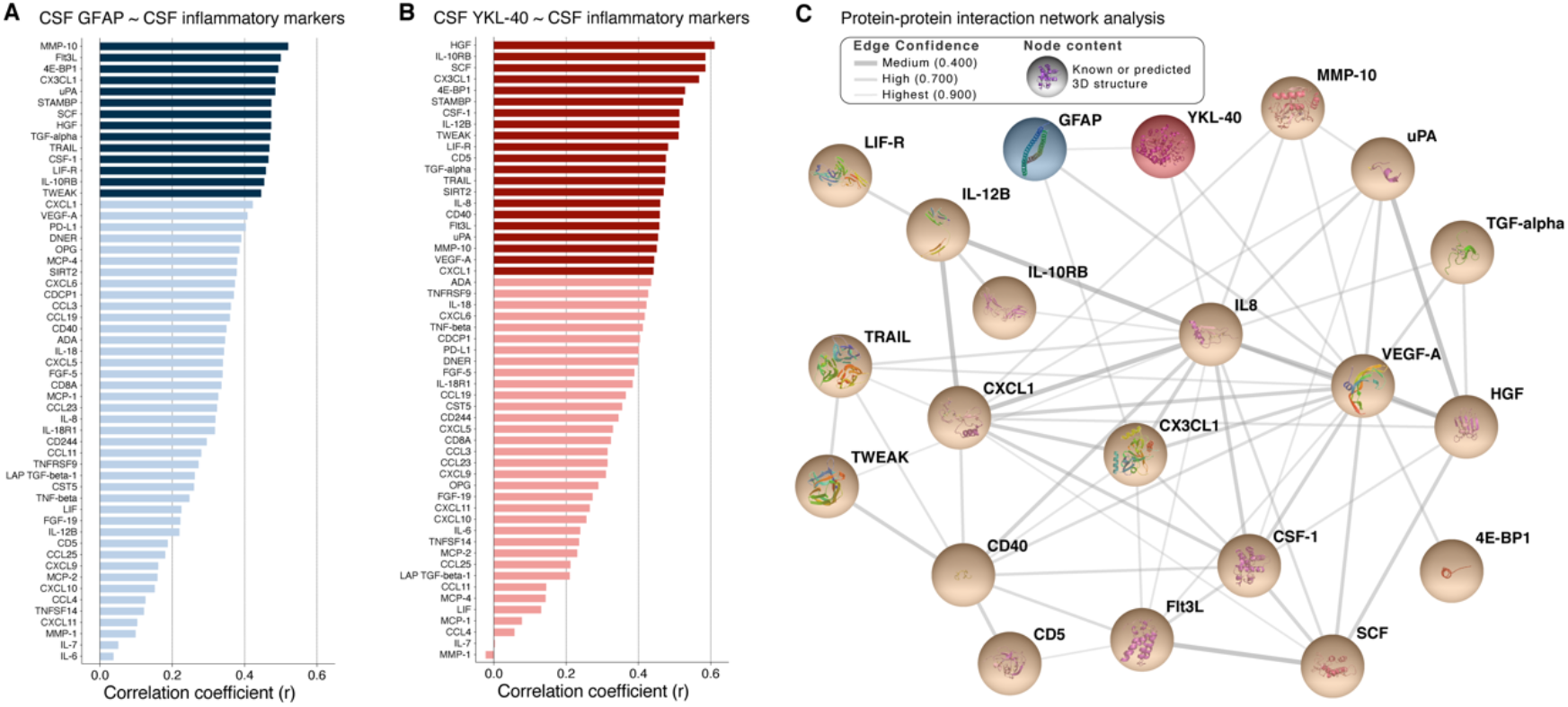
GFAP and YKL-40 are associated with CSF inflammatory makers. **(A)** The graph shows the correlations coefficients from age-adjusted partial correlations between CSF GFAP and CSF inflammation-related proteins. Dark blue bars represent statistically significant correlations after Bonferroni correction for multiple comparisons, whereas light blue bars represent correlations that did not survive Bonferroni correction. **(B)** The graph shows the correlations coefficients from age-adjusted partial correlations between CSF YKL-40 and CSF inflammation-related proteins. Dark red bars represent statistically significant correlations after Bonferroni correction for multiple comparisons, whereas light red bars represent correlations that did not survive Bonferroni correction. (**C**) Protein-protein interaction network analysis further support that GFAP and YKL-40 were inter-connected with several inflammation-related proteins. STAMBP and SIRT2 were disconnected to the other proteins in the network analysis; thus, these proteins were not represented in the figure. Abbreviations of inflammation-related proteins are reported in **Supplementary Table 5**.

## Discussion

Our results suggest that the two most widely used reactive astrocyte biomarkers, CSF GFAP and YKL-40, are differently associated with AD pathophysiological hallmarks in living humans. While GFAP levels increase in response to Aβ pathology, YKL-40 levels increase in response to tau pathology (**Fig. 4)**. We demonstrated that these reactive astrocyte biomarkers mediate the effects of Aβ and tau pathologies on hippocampal atrophy and cognitive impairment. Furthermore, we found that CSF GFAP and YKL-40 levels were closely related to the levels of CSF inflammation-related proteins.

**Figure 4.**
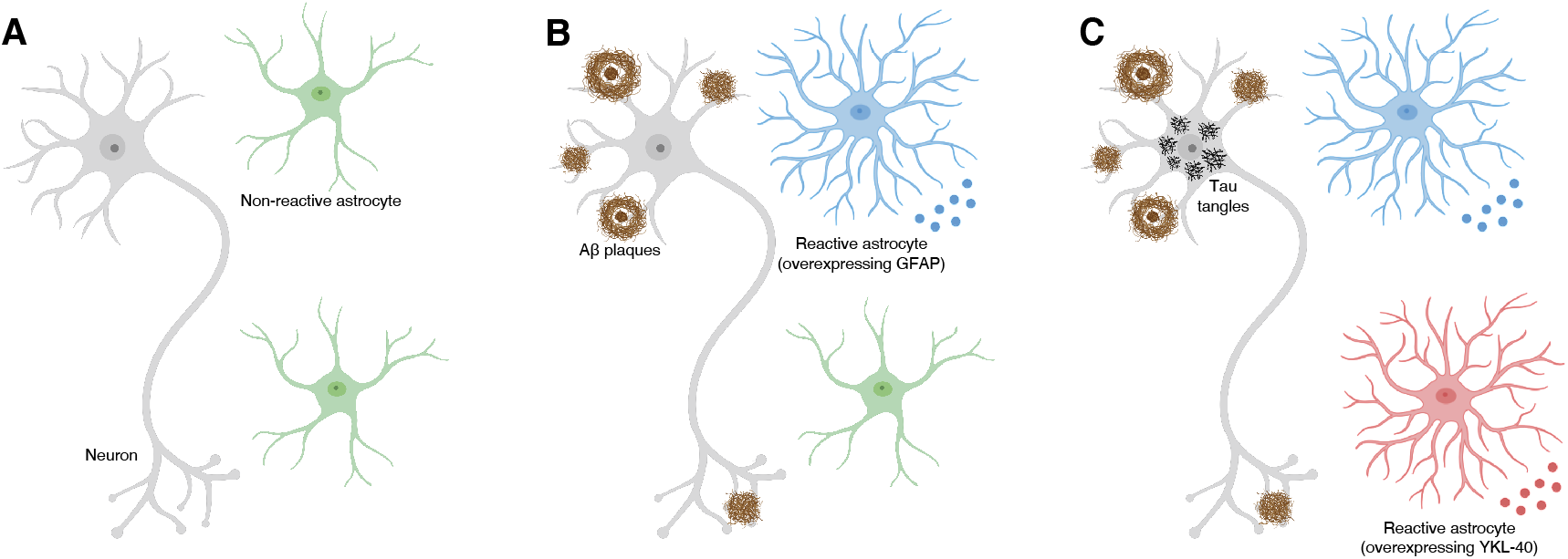
Schematic representation of reactive astrocyte signatures in the presence of brain Aβ and tau pathologies in AD. (**A**) In the absence of pathological stimuli, astrocytes are not reactive. (**B**) In response to brain Aβ accumulation, reactive astrocytes mainly overexpress GFAP. (**C**) In response to tau tangles accumulation, reactive astrocytes mainly overexpress YKL-40. Figure created with BioRender.com.

In the present study, CSF GFAP levels were associated with Aβ burden in typical AD brain regions, whereas no association was found with tau tangles accumulation. Importantly, we replicated these findings using plasma GFAP, which has been suggested to outperform CSF GFAP in the early detection of Aβ pathology^15^. It has already been reported that astrocytes assume multiple reactive phenotypes, overexpressing specific proteins depending on the pathological stimuli^8,10^. Previous *postmortem* observations suggested that reactive astrocytes overexpressing GFAP are found in the vicinity of Aβ plaques^1^. Furthermore, it was reported that the topography of GFAP-immunopositive astrocytes resembles the distribution of Aβ plaques in AD^20^. From the perspective of fluid biomarkers, GFAP levels are elevated in individuals within the AD spectrum^11,14,15,21-23^ and highly correlate with Aβ markers^11,14-16,21-23^. Altogether, these findings support the notion that astrocytes overexpress GFAP in response to brain Aβ deposition in AD. Even though YKL-40 is involved in the activation of innate immune cells, its function in the CNS remains poorly understood^8,10^. Importantly, the expression of this protein in the brain tissue has been detected in astrocytes, microglial cells, and infiltrating macrophages^24-26^. However, recent pathological evidence showed that YKL-40 strongly colocalizes with GFAP (astrocyte marker) but not with MAP2 (neuronal marker) and IBA-1 (microglia marker) in the AD brain^27^. Furthermore, another investigation reported that YKL-40 immunoreactivity was mainly observed within astrocytes but not within microglial cells in the frontal cortex of AD patients^28^. Thus, CSF YKL-40 is being increasingly accepted as a reactive astrocyte biomarker in AD^10,11^.

We observed that CSF YKL-40 levels were associated with tau but not Aβ pathology, indicating that YKL-40 levels in the CSF increase in response to tau tangles deposition in AD. *In vivo* studies suggest that CSF YKL-40 levels are elevated in AD^11,28^ and other tauopathies^17,29,30^, as well as correlate with CSF tau levels^16-19^. Accordingly, recent *postmortem* studies reported astrocyte overexpression of YKL-40 in AD and non-AD tauopathies *(e*.*g*., Pick’s disease, corticobasal degeneration, and progressive supranuclear palsy)^27^. Here, we expanded the aforementioned evidence by demonstrating that CSF YKL-40 levels were associated with tau accumulation -but not Aβ - in brain regions typically affected by AD-related tau pathology. Our models also showed that the association of CSF YKL-40 and tau pathology did not depend on plasma or CSF GFAP levels. Altogether, these results support the notion that astrocytes overexpress YKL-40 in response to tau tangles accumulation in AD.

We showed that reactive astrocyte biomarkers mediate the effect of Aβ and tau on neurodegeneration and cognitive impairment. Previous experimental studies demonstrated that reactive astrocytes actively promote neuronal injury^4,31-34^. Additionally, neuropathological evidence suggests that reactive astrogliosis is more prominent in brain regions more affected by degeneration in AD and other neurodegenerative conditions (*e*.*g*., Parkinson’s disease, Huntington’s disease, and multiple sclerosis)^4^. These findings further support the association of astrocyte reactivity with neurodegeneration and cognitive disfunction found in our study.

We found that CSF GFAP and YKL-40 levels were highly associated with several inflammatory markers. Protein-protein interaction network analysis revealed that CSF GFAP and YKL-40 were closely related to neuroinflammatory proteins previously associated with AD progression such as IL-8^35^, CX3CL1^36^, CXCL1^37^, MMP-10^38^, TRAIL^39^, HGF^40^, CSF-1^41^, and 4E-BP1^42^, which supports a close link between astrocyte reactivity and neuroinflammation. These findings corroborate that astrocyte reactivity can be triggered by inflammatory mediators released by activated microglia^4^. Moreover, recent evidence from animal models and *post-mortem* brain tissue demonstrated that Aβ and tau pathologies induce molecular astrocyte signatures associated with the activation of inflammatory pathways^9^. These results suggest that astrocyte reactivity might be an important component of the complex interplay between brain proteinopathies and neuroinflammation in AD.

Our findings have important implications in the context of emerging anti-Aβ and anti-tau therapies for AD^43,44^. Given that AD is a complex and multifaceted disease^10^, it is reasonable to postulate that the combination of therapies - rather than single-target treatments - can offer more effective outcomes. A recent consensus reinforced the importance of developing astrocyte-based therapies for neurodegenerative conditions, suggesting that studies focusing on the characterization of *in vivo* astrocyte biomarkers should be a priority in order to achieve this goal^8^. In line with this, our results suggest that drugs targeting specific reactive astrocyte phenotypes could potentially be used in combination with anti-Aβ and anti-tau therapies to enhance treatment response in future disease-modifying clinical trials.

This study has methodological limitations. Our results demonstrate astrocyte biomarker signatures associated with Aβ and tau deposition in living patients. It is possible that subjects presenting similar astrocyte biomarker signatures have different astrocyte molecular, morphological, and functional characteristics^8^, which was not evaluated in the current investigation. Given that we used fluid levels of GFAP and YKL-40, it is also important to acknowledge the lack of topographical information provided by these biomarkers to assess reactive astrogliosis. Our cohort is composed of individuals that are motivated to participate in a dementia study, potentially being a source of self-selection bias. Additionally, we used a cross-sectional design and modeled AD progressions using individuals across the disease spectrum. It would be highly desirable to replicate our findings using a longitudinal design with multiple time points to better characterize the sequential relation of markers.

To conclude, we observed that plasma and CSF GFAP levels associate with Aβ while CSF YKL-40 levels associate with tau pathology, suggesting the existence of astrocyte biomarker signatures of Aβ and tau tangles in the living AD brain.

## Materials and Methods

### Participants

Study participants are part of the Translational Biomarkers in Aging and Dementia (TRIAD) cohort, McGill University, Canada (https://triad.tnl-mcgill.com). Participants from the community or outpatients at the McGill University Research Centre for Studies in Aging were recruited through different sources, such as printed materials, word of mouth, and referrals. Exclusion criteria included inability to speak English or French, inadequate visual and auditory capacities for neuropsychologic assessment, active substance abuse, major surgery, recent head trauma, medical contraindication for PET or magnetic resonance imaging (MRI), currently being enrolled in other studies, and neurological, psychiatric, or systemic comorbidities that were not adequately treated with a stable medication regimen. The Douglas Mental Health University Institute Research Ethics Board and the Montreal Neurological Instituted PET working committee approved this study. All participants provided written informed consent.

We assessed 75 CU and 46 CI participants with 50 years of age or older. In addition to CSF GFAP and YKL-40, individuals had available Aβ-PET, tau-PET, and MRI at the same visit, as well as *APOE* genotyping. The mean time difference between CSF collection and imaging was 0.6 months (range: 0 – 11.7 months). Two outliers were detected [CSF GFAP levels that were three standard deviations (SD) above the mean of the whole population] and excluded from subsequent analyses, as previously done^45,46^. For a detailed description of the selection of study participants, see **Supplementary Fig. 3**. All individuals had detailed neuropsychological testing including MMSE and Clinical Dementia Rating (CDR). CU subjects had a CDR of 0 and no objective cognitive impairment. MCI patients had CDR of 0.5, subjective and objective cognitive impairments, and preserved activities of daily living^47^. Mild-to-moderate AD dementia patients had CDR of between 0.5 and 2 and met the National Institute on Aging and the Alzheimer’s Association (NIA-AA) criteria for probable AD^48^. In accordance with the updated 2018 NIA-AA Research Framework^49^, AD dementia participants were required to be Aβ positive, similar to previous publications^50,51^.

### Fluid biomarkers

All samples were analyzed at the Clinical Neurochemistry Laboratory at the University of Gothenburg, Sweden. CSF and plasma GFAP were quantified using a commercial single-plex assay (No. 102336) on the Single molecule array (Simoa) HD-X (Quanterix, Billerica, Maryland, USA)^15^. CSF YKL-40 was measured using a commercial ELISA assay (R&D Systems, Minneapolis, USA)^15^. Moreover, a subset of 62 participants had CSF samples analyzed using a multiplex immunoassay for a panel of 92 proteins related to inflammatory diseases and associated biological processes (Olink; https://www.olink.com/products/inflammation/). We excluded 37 markers with a high percentage (>15%) of values below the lower detection limit, as previously described^52^. The complete list of abbreviations for inflammation-related markers is found in **Supplementary Table 5**.

### Neuroimaging biomarkers

T_1_-weighted MRIs were acquired on a 3T Siemens Magnetom using a standard head coil and the magnetization prepared rapid acquisition gradient echo (MPRAGE) sequence was used to obtain high-resolution structural images of the whole brain. Aβ-PET ([^18^F]AZD4694; 40–70 min post-injection) and tau-PET ([^18^F]MK-6240; 90–110 min post-injection) scans were acquired on a Siemens high-resolution research tomograph. Radiosynthesis of PET tracers have been described elsewhere^53,54^. Aβ-PET and tau-PET scans were reconstructed using the ordered subset expectation maximization algorithm on a 4D volume with three frames (3 × 600 seconds) and four frames (4 × 300 seconds), respectively^53^. Details regarding MRI and PET acquisition and processing are described in the **Supplementary Methods**.

We used the Desikan-Killiany-Tourville atlas to define ROIs^55^. For Aβ-PET, a global neocortical SUVR was estimated from the following brain regions: precuneus, prefrontal, orbitofrontal, parietal, temporal, and cingulate cortices^56^. Aβ (A) positivity was defined as neocortical Aβ-PET SUVR ≥ 1.55 following a published threshold for [^18^F]AZD4694 Aβ-PET^57^. For tau-PET, a temporal meta-ROI SUVR was estimated from the following brain regions: entorhinal, hippocampus, fusiform, parahippocampal, inferior temporal, and middle temporal^56^. Tau (T) positivity was defined as temporal meta-ROI tau-PET SUVR ≥ 1.24 as described elsewhere^58^. Of note, there were no A-T+ subjects as we did not include Aβ negative individuals with a clinical diagnosis of AD dementia. Hippocampal volume was adjusted for total intracranial volume^59^.

### Statistical analysis

Statistical analyses were conducted in the R free software (version 4.0.2, http://www.r-project.org/) for non-imaging analyses, and MATLAB software (version 9.2, http://www.mathworks.com) with VoxelStats package for imaging analyses^60^. Student’s t test (continuous variables) and contingency χ2 test (categorical variables) tested demographic differences. Spearman rank test was used to assess the correlations between reactive astrocyte biomarkers. Analyses of variance with Tukey’s multiple comparisons test was used to compare adjusted levels of GFAP and YKL-40 markers across groups defined based on Aβ-PET and tau-PET status. Adjusted values were the residuals of the regressions between biomarker level and covariates of interest (age, sex, and *APOE* ε4 status). The associations of Aβ-PET and tau-PET with reactive astrocyte biomarkers were tested using ROI-based linear regressions, as well as voxel-wise linear regressions. Models were adjusted for age, sex, cognitive status, and *APOE* ε4 status. In ROI-based multiple regression models, partial residuals generated with the R function termplot were used to graphically represent the investigated associations^61,62^. In voxel-wise analyses, multiple comparisons correction was performed using random field theory (RFT)^63^, with a voxel threshold of *P* < 0.001. We evaluated whether CSF GFAP and YKL-40 mediate the effect of Aβ and tau pathologies on neurodegeneration and cognition using structural equation modeling, R package “lavaan”^64^. The fit of the structural equation models was classified as good if: RMSEA < 0.05 (acceptable 0.05 - 0.08); CFI > 0.97 (acceptable: 0.95 - 0.97); and SRMR < 0.05 (acceptable: 0.05 - 0.10)^65,66^. The association of CSF GFAP and YKL-40 with CSF inflammation-related proteins was assessed with age-adjusted partial correlations with Bonferroni correction for multiple comparisons. Furthermore, the Search Tool for the Retrieval of Interacting Genes/Proteins (STRING) database (version 11.5)^67^ was used to construct a protein-protein interaction network including GFAP and YKL-40, as well as the inflammation-related proteins that were significantly correlated to these reactive astrocyte biomarkers. For graphical representation, we filtered connections with STRING confidence interaction scores > 0.4. The statistical significance level was set as *P* < 0.05, two-tailed. Continuous variables were standardized before model entry in ROI-based linear regressions and structural equation models to facilitate comparison across estimates.

## Data Availability

The data from the Translational Biomarkers in Aging and Dementia (TRIAD) study will be made available from the senior authors upon reasonable request. Such arrangements are subject to standard data-sharing agreements. Of note, the data used in the present work is not publicly available because the information could compromise the participants' privacy.

## Data availability

The data from the TRIAD study will be made available from the senior authors upon reasonable request. Such arrangements are subject to standard data-sharing agreements. Of note, the data used in the present work is not publicly available because the information could compromise the participants’ privacy.

## Acknowledgments

We acknowledge all study participants and the staff of the McGill Center for studies in Aging. Cerveau Technologies enabled the use of [^18^F]MK-6240. We would also like to thank Dean Jolly, Alexey Kostikov, Monica Samoila-Lactatus, Karen Ross, Mehdi Boudjemeline, and Sandy Li for assist in the radiochemistry production, as well as Richard Strauss, Edith Strauss, Guylaine Gagne, Carley Mayhew, Tasha Vinet-Celluci, Karen Wan, Sarah Sbeiti, Meong Jin Joung, Miloudza Olmand, Rim Nazar, Hung-Hsin Hsiao, Reda Bouhachi, and Arturo Aliaga for consenting subjects and/or helping with the acquisition of the data.

## Author contributions

J.P.F-S., E.R.Z., P.R-N., and T.A.P. conceived the study. J.P.F-S., E.R.Z., P.R-N., and T.A.P. prepared the prepared figures, tables, and drafted the manuscript with input from P.C.L.F., B.B., C.T., Y-T.W., D.T.L., W.S.B., A.L.B., N.J.A., M.A.D.B., A.R., J.T., F.Z.L., M.C., S.S., G.B., M.S.K., J.S., N.R., V.P., N.M.P., W.E.K., D.L.T., A.C., V.L.V., S.G., K.B., H.Z., D.O.S., and T.K.K. J.P.F-S., P.C.L.F., B.B., C.T., Y-T.W., D.T.L., W.S.B., A.L.B., N.J.A., M.A.D.B., A.R., J.T., F.Z.L., M.C., S.S., G.B., M.S.K., J.S., N.R., V.P., E.R.Z., P.R-N., and T.A.P. performed the acquisitions, processing, quality control, and/or interpretation of the data. T.A.P., P.R-N., and E.R.Z. supervised this work. M.A.D.B. and B.B. assisted in the protein-protein interaction network analysis. D.L.T. assisted in the statistical analysis. All authors revised and approved the final paper draft.

## References

1 Serrano-Pozo, A. et al. Reactive glia not only associates with plaques but also parallels tangles in Alzheimer’s disease. Am J Pathol 179, 1373–1384, doi:10.1016/j.ajpath.2011.05.047 (2011).

2 Marutle, A. et al. (3)H-deprenyl and (3)H-PIB autoradiography show different laminar distributions of astroglia and fibrillar beta-amyloid in Alzheimer brain. J Neuroinflammation 10, 90, doi:10.1186/1742-2094-10-90 (2013).

3 Lemoine, L., Saint-Aubert, L., Nennesmo, I., Gillberg, P. G. & Nordberg, A. Cortical laminar tau deposits and activated astrocytes in Alzheimer’s disease visualised by (3)H-THK5117 and (3)H-deprenyl autoradiography. Sci Rep 7, 45496, doi:10.1038/srep45496 (2017).

4 Liddelow, S. A. et al. Neurotoxic reactive astrocytes are induced by activated microglia. Nature 541, 481–487, doi:10.1038/nature21029 (2017).

5 Habib, N. et al. Disease-associated astrocytes in Alzheimer’s disease and aging. Nat Neurosci 23, 701–706, doi:10.1038/s41593-020-0624-8 (2020).

6 Perez-Nievas, B. G. & Serrano-Pozo, A. Deciphering the Astrocyte Reaction in Alzheimer’s Disease. Front Aging Neurosci 10, 114, doi:10.3389/fnagi.2018.00114 (2018).

7 Galea, E. et al. Multi-transcriptomic analysis points to early organelle dysfunction in human astrocytes in Alzheimer’s disease. medRxiv, doi:10.1101/2021.02.25.21252422 (2021).

8 Escartin, C. et al. Reactive astrocyte nomenclature, definitions, and future directions. Nat Neurosci 24, 312–325, doi:10.1038/s41593-020-00783-4 (2021).

9 Jiwaji, Z. et al. Reactive astrocytes acquire neuroprotective as well as deleterious signatures in response to Tau and Ass pathology. Nat Commun 13, 135, doi:10.1038/s41467-021-27702-w (2022).

10 Carter, S. F. et al. Astrocyte Biomarkers in Alzheimer’s Disease. Trends Mol Med 25, 77–95, doi:10.1016/j.molmed.2018.11.006 (2019).

11 Bellaver, B. et al. Astrocyte Biomarkers in Alzheimer Disease: A Systematic Review and Meta-analysis. Neurology, doi:10.1212/WNL.0000000000012109 (2021).

12 Sun, M. et al. A candidate biomarker of glial fibrillary acidic protein in CSF and blood in differentiating multiple sclerosis and its subtypes: A systematic review and meta-analysis. Mult Scler Relat Disord 51, 102870, doi:10.1016/j.msard.2021.102870 (2021).

13 Burman, J. et al. YKL-40 is a CSF biomarker of intrathecal inflammation in secondary progressive multiple sclerosis. J Neuroimmunol 292, 52–57, doi:10.1016/j.jneuroim.2016.01.013 (2016).

14 Pereira, J. B. et al. Plasma GFAP is an early marker of amyloid-beta but not tau pathology in Alzheimer’s disease. Brain, doi:10.1093/brain/awab223 (2021).

15 Benedet, A. L. et al. Differences Between Plasma and Cerebrospinal Fluid Glial Fibrillary Acidic Protein Levels Across the Alzheimer Disease Continuum. JAMA Neurol, doi:10.1001/jamaneurol.2021.3671 (2021).

16 Mila-Aloma, M. et al. Amyloid beta, tau, synaptic, neurodegeneration, and glial biomarkers in the preclinical stage of the Alzheimer’s continuum. Alzheimers Dement 16, 1358–1371, doi:10.1002/alz.12131 (2020).

17 Alcolea, D. et al. Relationship between beta-Secretase, inflammation and core cerebrospinal fluid biomarkers for Alzheimer’s disease. J Alzheimers Dis 42, 157–167, doi:10.3233/JAD-140240 (2014).

18 Alcolea, D. et al. Amyloid precursor protein metabolism and inflammation markers in preclinical Alzheimer disease. Neurology 85, 626–633, doi:10.1212/WNL.0000000000001859 (2015).

19 Antonell, A. et al. Cerebrospinal fluid level of YKL-40 protein in preclinical and prodromal Alzheimer’s disease. J Alzheimers Dis 42, 901–908, doi:10.3233/JAD-140624 (2014).

20 Beach, T. G. & McGeer, E. G. Lamina-specific arrangement of astrocytic gliosis and senile plaques in Alzheimer’s disease visual cortex. Brain Res 463, 357–361, doi:10.1016/0006-8993(88)90410-6 (1988).

21 Oeckl, P. et al. Glial Fibrillary Acidic Protein in Serum is Increased in Alzheimer’s Disease and Correlates with Cognitive Impairment. J Alzheimers Dis 67, 481–488, doi:10.3233/JAD-180325 (2019).

22 Simren, J. et al. The diagnostic and prognostic capabilities of plasma biomarkers in Alzheimer’s disease. Alzheimers Dement 17, 1145–1156, doi:10.1002/alz.12283 (2021).

23 Schulz, I. et al. Systematic Assessment of 10 Biomarker Candidates Focusing on alpha-Synuclein-Related Disorders. Mov Disord, doi:10.1002/mds.28738 (2021).

24 Hinsinger, G. et al. Chitinase 3-like proteins as diagnostic and prognostic biomarkers of multiple sclerosis. Mult Scler 21, 1251–1261, doi:10.1177/1352458514561906 (2015).

25 Canto, E. et al. Chitinase 3-like 1: prognostic biomarker in clinically isolated syndromes. Brain 138, 918–931, doi:10.1093/brain/awv017 (2015).

26 Dichev, V., Kazakova, M. & Sarafian, V. YKL-40 and neuron-specific enolase in neurodegeneration and neuroinflammation. Rev Neurosci 31, 539–553, doi:10.1515/revneuro-2019-0100 (2020).

27 Querol-Vilaseca, M. et al. YKL-40 (Chitinase 3-like I) is expressed in a subset of astrocytes in Alzheimer’s disease and other tauopathies. J Neuroinflammation 14, 118, doi:10.1186/s12974-017-0893-7 (2017).

28 Craig-Schapiro, R. et al. YKL-40: a novel prognostic fluid biomarker for preclinical Alzheimer’s disease. Biol Psychiatry 68, 903–912, doi:10.1016/j.biopsych.2010.08.025 (2010).

29 Teunissen, C. E. et al. Novel diagnostic cerebrospinal fluid biomarkers for pathologic subtypes of frontotemporal dementia identified by proteomics. Alzheimers Dement (Amst) 2, 86–94, doi:10.1016/j.dadm.2015.12.004 (2016).

30 Alcolea, D. et al. CSF sAPPbeta, YKL-40, and neurofilament light in frontotemporal lobar degeneration. Neurology 89, 178–188, doi:10.1212/WNL.0000000000004088 (2017).

31 Liddelow, S. A. & Barres, B. A. Reactive Astrocytes: Production, Function, and Therapeutic Potential. Immunity 46, 957–967, doi:10.1016/j.immuni.2017.06.006 (2017).

32 Guttenplan, K. A. et al. Neurotoxic Reactive Astrocytes Drive Neuronal Death after Retinal Injury. Cell Rep 31, 107776, doi:10.1016/j.celrep.2020.107776 (2020).

33 Guttenplan, K. A. et al. Knockout of reactive astrocyte activating factors slows disease progression in an ALS mouse model. Nat Commun 11, 3753, doi:10.1038/s41467-020-17514-9 (2020).

34 Guttenplan, K. A. et al. Neurotoxic reactive astrocytes induce cell death via saturated lipids. Nature, doi:10.1038/s41586-021-03960-y (2021).

35 McLarnon, J. G. Chemokine Interleukin-8 (IL-8) in Alzheimer’s and Other Neurodegenerative Diseases. Journal of Alzheimer’s Disease & Parkinsonism 6, doi:10.4172/2161-0460.1000273 (2016).

36 Bolos, M. et al. Absence of CX3CR1 impairs the internalization of Tau by microglia. Mol Neurodegener 12, 59, doi:10.1186/s13024-017-0200-1 (2017).

37 Zhang, K. et al. CXCL1 contributes to beta-amyloid-induced transendothelial migration of monocytes in Alzheimer’s disease. PLoS One 8, e72744, doi:10.1371/journal.pone.0072744 (2013).

38 Duits, F. H. et al. Matrix Metalloproteinases in Alzheimer’s Disease and Concurrent Cerebral Microbleeds. J Alzheimers Dis 48, 711–720, doi:10.3233/JAD-143186 (2015).

39 Cantarella, G. et al. Neutralization of TNFSF10 ameliorates functional outcome in a murine model of Alzheimer’s disease. Brain 138, 203–216, doi:10.1093/brain/awu318 (2015).

40 Fenton, H. et al. Hepatocyte growth factor (HGF/SF) in Alzheimer’s disease. Brain Res 779, 262–270, doi:10.1016/s0006-8993(97)00958-x (1998).

41 Murphy, G. M., Jr., Zhao, F., Yang, L. & Cordell, B. Expression of macrophage colony-stimulating factor receptor is increased in the AbetaPP(V717F) transgenic mouse model of Alzheimer’s disease. Am J Pathol 157, 895–904, doi:10.1016/s0002-9440(10)64603-2 (2000).

42 Li, X., Alafuzoff, I., Soininen, H., Winblad, B. & Pei, J. J. Levels of mTOR and its downstream targets 4E-BP1, eEF2, and eEF2 kinase in relationships with tau in Alzheimer’s disease brain. FEBS J 272, 4211–4220, doi:10.1111/j.1742-4658.2005.04833.x (2005).

43 Cavazzoni, P. FDA’s Decision to Approve New Treatment for Alzheimer’s Disease, <https://www.fda.gov/drugs/news-events-human-drugs/fdas-decision-approve-new-treatment-alzheimers-disease> (2021).

44 Cummings, J., Lee, G., Zhong, K., Fonseca, J. & Taghva, K. Alzheimer’s disease drug development pipeline: 2021. Alzheimers Dement (N Y) 7, e12179, doi:10.1002/trc2.12179 (2021).

45 Karikari, T. K. et al. Diagnostic performance and prediction of clinical progression of plasma phospho-tau181 in the Alzheimer’s Disease Neuroimaging Initiative. Mol Psychiatry 26, 429–442, doi:10.1038/s41380-020-00923-z (2021).

46 Mattsson-Carlgren, N. et al. Longitudinal plasma p-tau217 is increased in early stages of Alzheimer’s disease. Brain 143, 3234–3241, doi:10.1093/brain/awaa286 (2020).

47 Petersen, R. C. Mild cognitive impairment as a diagnostic entity. J Intern Med 256, 183–194, doi:10.1111/j.1365-2796.2004.01388.x (2004).

48 McKhann, G. M. et al. The diagnosis of dementia due to Alzheimer’s disease: recommendations from the National Institute on Aging-Alzheimer’s Association workgroups on diagnostic guidelines for Alzheimer’s disease. Alzheimers Dement 7, 263–269, doi:10.1016/j.jalz.2011.03.005 (2011).

49 Jack, C. R., Jr. et al. NIA-AA Research Framework: Toward a biological definition of Alzheimer’s disease. Alzheimers Dement 14, 535–562, doi:10.1016/j.jalz.2018.02.018 (2018).

50 Ossenkoppele, R. et al. Discriminative Accuracy of [18F]flortaucipir Positron Emission Tomography for Alzheimer Disease vs Other Neurodegenerative Disorders. JAMA 320, 1151–1162, doi:10.1001/jama.2018.12917 (2018).

51 Palmqvist, S. et al. Discriminative Accuracy of Plasma Phospho-tau217 for Alzheimer Disease vs Other Neurodegenerative Disorders. JAMA 324, 772–781, doi:10.1001/jama.2020.12134 (2020).

52 Pascoal, T. A. et al. Microglial activation and tau propagate jointly across Braak stages. Nat Med 27, 1592–1599, doi:10.1038/s41591-021-01456-w (2021).

53 Pascoal, T. A. et al. In vivo quantification of neurofibrillary tangles with [(18)F]MK-6240. Alzheimers Res Ther 10, 74, doi:10.1186/s13195-018-0402-y (2018).

54 Cselenyi, Z. et al. Clinical validation of 18F-AZD4694, an amyloid-beta-specific PET radioligand. J Nucl Med 53, 415–424, doi:10.2967/jnumed.111.094029 (2012).

55 Klein, A. & Tourville, J. 101 labeled brain images and a consistent human cortical labeling protocol. Front Neurosci 6, 171, doi:10.3389/fnins.2012.00171 (2012).

56 Jack, C. R., Jr. et al. Defining imaging biomarker cut points for brain aging and Alzheimer’s disease. Alzheimers Dement 13, 205–216, doi:10.1016/j.jalz.2016.08.005 (2017).

57 Therriault, J. et al. Determining Amyloid-beta Positivity Using (18)F-AZD4694 PET Imaging. J Nucl Med 62, 247–252, doi:10.2967/jnumed.120.245209 (2021).

58 Therriault, J. et al. Frequency of Biologically Defined Alzheimer Disease in Relation to Age, Sex, APOE epsilon4, and Cognitive Impairment. Neurology 96, e975–e985, doi:10.1212/WNL.0000000000011416 (2021).

59 Hansen, T. I., Brezova, V., Eikenes, L., Haberg, A. & Vangberg, T. R. How Does the Accuracy of Intracranial Volume Measurements Affect Normalized Brain Volumes? Sample Size Estimates Based on 966 Subjects from the HUNT MRI Cohort. AJNR Am J Neuroradiol 36, 1450–1456, doi:10.3174/ajnr.A4299 (2015).

60 Mathotaarachchi, S. et al. VoxelStats: A MATLAB Package for Multi-Modal Voxel-Wise Brain Image Analysis. Front Neuroinform 10, 20, doi:10.3389/fninf.2016.00020 (2016).

61 Larsen, W. A. & McCleary, S. J. The Use of Partial Residual Plots in Regression Analysis. Technometrics 14, 781–790, doi:10.1080/00401706.1972.10488966 (1972).

62 Ryan, T. P. Modern Regression Methods. (John Wiley & Sons, Inc, 2008).

63 Worsley, K. J., Taylor, J. E., Tomaiuolo, F. & Lerch, J. Unified univariate and multivariate random field theory. Neuroimage 23 Suppl 1, S189–195, doi:10.1016/j.neuroimage.2004.07.026 (2004).

64 Rosseel, Y. lavaan: An R Package for Structural Equation Modeling. Journal of Statistical Software 48, 1–36, doi:10.18637/jss.v048.i02 (2012).

65 Mueller, R. O. & Hancock, G. R. in Best Practices in Quantitative Methods (ed J. Osborne) Ch. 32, 488-508 (SAGE, 2008).

66 Schermelleh-Engel, K., Moosbrugger, H. & Müller, H. Evaluating the Fit of Structural Equation Models: Tests of Significance and Descriptive Goodness-of-Fit Measures. Methods of Psychological Research 8, 23–74 (2003).

67 Szklarczyk, D. et al. STRING v11: protein-protein association networks with increased coverage, supporting functional discovery in genome-wide experimental datasets. Nucleic Acids Res 47, D607–D613, doi:10.1093/nar/gky1131 (2019).

