## Supplementary Information for "Astrocyte biomarker signatures of amyloid-β and tau pathologies in Alzheimer’s disease"

Ferrari-Souza *et al.*

**Supplementary Methods.** MRI and PET acquisition and processing.

**Supplementary Figure 1.** Correlation between reactive astrocyte biomarkers.

**Supplementary Figure 2.** Sensitivity analyses testing the associations of Aβ-PET and tau-PET with reactive astrocyte biomarkers using plasma GFAP instead of CSF GFAP.

**Supplementary Figure 3.** Flowchart of included participants.

**Supplementary Table 1.** Associations of CSF GFAP and YKL-40 with neocortical Aβ-PET and temporal meta-ROI tau-PET.

**Supplementary Table 2.** Structural equation model coefficients and associated statistics for Figure 2A.

**Supplementary Table 3.** Structural equation model coefficients and associated statistics for Figure 2B.

**Supplementary Table 4.** Demographics of the subset of individuals with available CSF inflammation-related proteins.

**Supplementary Table 5.** Abbreviations of inflammation-related proteins.

Abbreviations: AD = Alzheimer's disease; ADAD = autosomal dominant Alzheimer's disease; ADNI = Alzheimer's Disease Neuroimaging Initiative; *APOE* ε4 = Apolipoprotein E ε4; Aβ = amyloid-β; CFI = comparative fit index; CSF = cerebrospinal fluid; CI = cognitively impaired; CU = cognitively unimpaired; GFAP = glial fibrillary acidic protein; MCI = mild cognitive impairment; MMSE = Mini-Mental State Examination; MPRAGE = magnetization prepared rapid acquisition gradient echo; MNI = Montreal Neurological Institute; NS = not significant; PET = positron emission tomography; RMSEA = root mean squared error of approximation; SD = standard deviation; SRMR = standardized root mean square residual; SUVR = standardized uptake value ratio; YKL-40 = chitinase-3-like protein 1.

**Supplementary Methods. MRI and PET acquisition and processing.**

Structural MRI was acquired at the MNI using a 3-T Siemens Magnetom using a standard head coil. We used the MPRAGE MRI (TR: 2300 ms, TE: 2.96ms) sequence to obtain high-resolution structural images of the whole brain (9° flip angle, coronal orientation perpendicular to the double spin echo sequence, 1x1 $mm^2$ in-plane resolution of 1 mm slab thickness). Then, the Statistical Parametric Mapping 12 segmentation tool was used to segment anatomical images into probabilistic grey matter and white matter maps. Each grey matter probability map was then non-linearly registered (with modulation) to the ADNI template using DARTEL[1] and voxel values were modulated by multiplying them by the Jacobian determinants derived from the spatial normalization step[2]. MRI images were smoothed using a Gaussian kernel of full width half maximum of 8 mm. Lastly, we visually inspected all images to ensure proper alignment to the ADNI template. Aβ-PET ([$^{18}$F]AZD4694; 40–70 min post-injection) and tau-PET ([$^{18}$F]MK-6240; 90–110 min post-injection) scans were acquired on a Siemens High Resolution Research Tomograph. Aβ-PET and tau-PET scans were reconstructed using the ordered subset expectation maximization algorithm on a 4D volume with three frames (3 x 600 seconds) and four frames (4 x 300 seconds), respectively[3]. Then, attenuation correction was performed using a 6-minute transmission scan with a rotating $^{137}$Cs point source. Furthermore, PET images were corrected for motion, dead time, decay, and scattered and random coincidences. Following an in-house pipeline, T1-weighted MRI data was corrected for non-uniformity and field distortion. Subsequently, linear co-registration and non-linear spatial normalization for the MNI template was performed through linear and non-linear transformation in two main steps: (i) PET registration to the correspondent T1-weighted MRI, and (ii) T1-weighted MRI registration to the MNI reference space. PET images were spatially smoothed to achieve a final resolution of 8 mm full width at half-maximum width. SUVRs were calculated using the whole cerebellum grey matters reference region for [$^{18}$F]AZD4694 Aβ-PET[4] and using inferior cerebellum as reference region for [$^{18}$F]MK-6240 tau-PET[5].

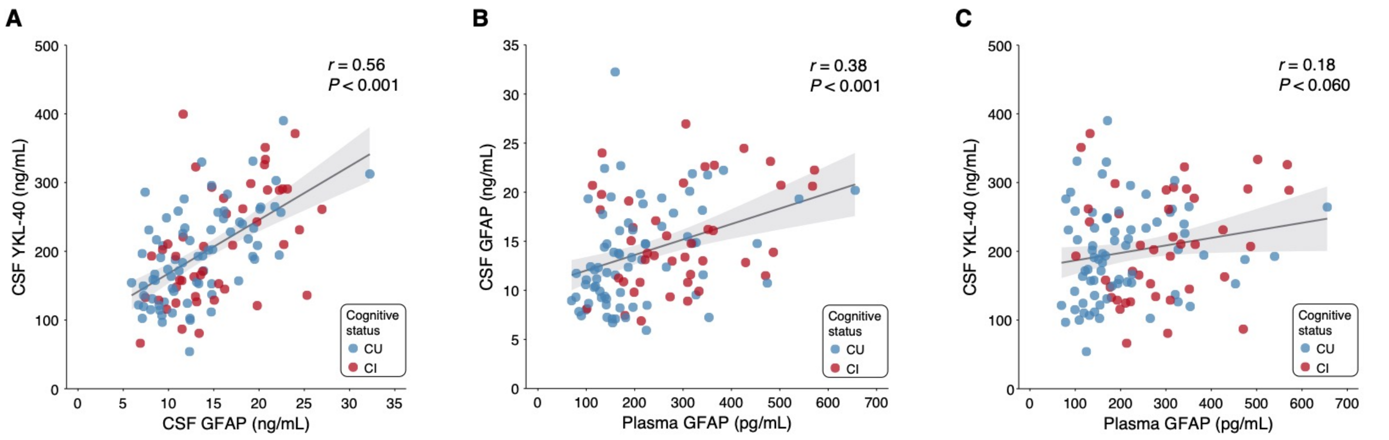


**Supplementary Figure 1. Correlation between reactive astrocyte biomarkers.** Scatter plot showing the results of Spearman's rank correlations between (**A**) CSF GFAP and CSF YKL-40, (**B**) plasma GFAP and CSF GFAP, and (**C**) plasma GFAP and CSF YKL-40. The error bands indicate the 95% confidence intervals. Individuals are colored by cognitive status. Noteworthy, analyses involving plasma GFAP were conducted in a subset of 114 individuals; from the total study population of 121 subjects, five participants did not have available plasma GFAP measures and two were excluded because they were considered outliers (plasma GFAP concentrations three SD above the mean of the population).

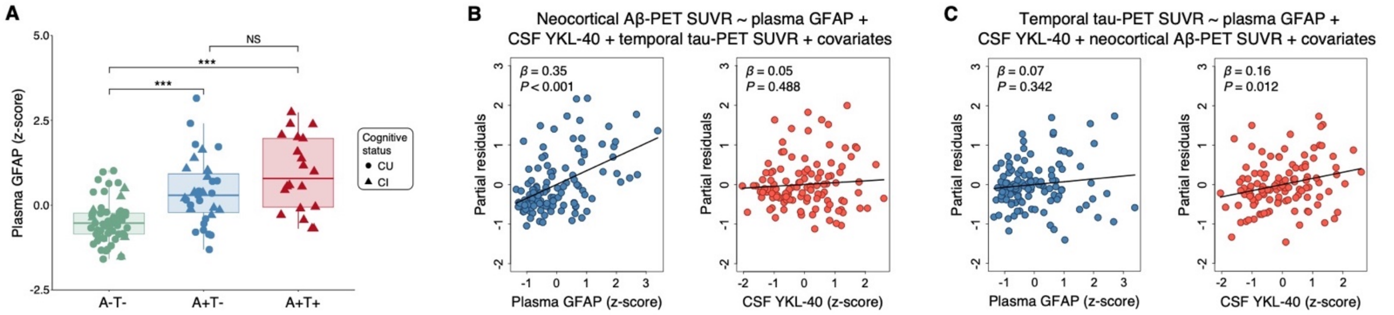


**Supplementary Figure 2. Sensitivity analyses testing the associations of Aβ-PET and tau-PET with reactive astrocyte biomarkers using plasma GFAP instead of CSF GFAP.** (**A**) Box-and-whisker plot of plasma GFAP levels adjusted for age-, sex-, and *APOE* ε4 status across AT groups. The horizontal line in each box represents the median; box ends represent the 25th and 75th percentiles. Groups were compared using analyses of variance with Tukey's multiple comparison test (**P*<0.05, ***P*<0.01, ****P*<0.001). (**B**) Partial residual plots of linear regressions testing the associations of neocortical Aβ-PET SUVR with plasma GFAP and CSF YKL-40 levels adjusting for temporal meta-ROI tau-PET SUVR, age, sex, cognitive status, and *APOE* ε4 status. (**C**) Partial residual plots of linear regressions testing the associations of temporal meta-ROI tau-PET SUVR with plasma GFAP and CSF YKL-40 levels adjusting for neocortical Aβ-PET SUVR, age, sex, cognitive status, and *APOE* ε4 status. Of note, analyses involving plasma GFAP were conducted in a subset of 114 individuals; from the total study population of 121 subjects, five participants did not have available plasma GFAP measures and two were excluded because they were considered outliers (plasma GFAP concentrations three SD above the mean of the population).

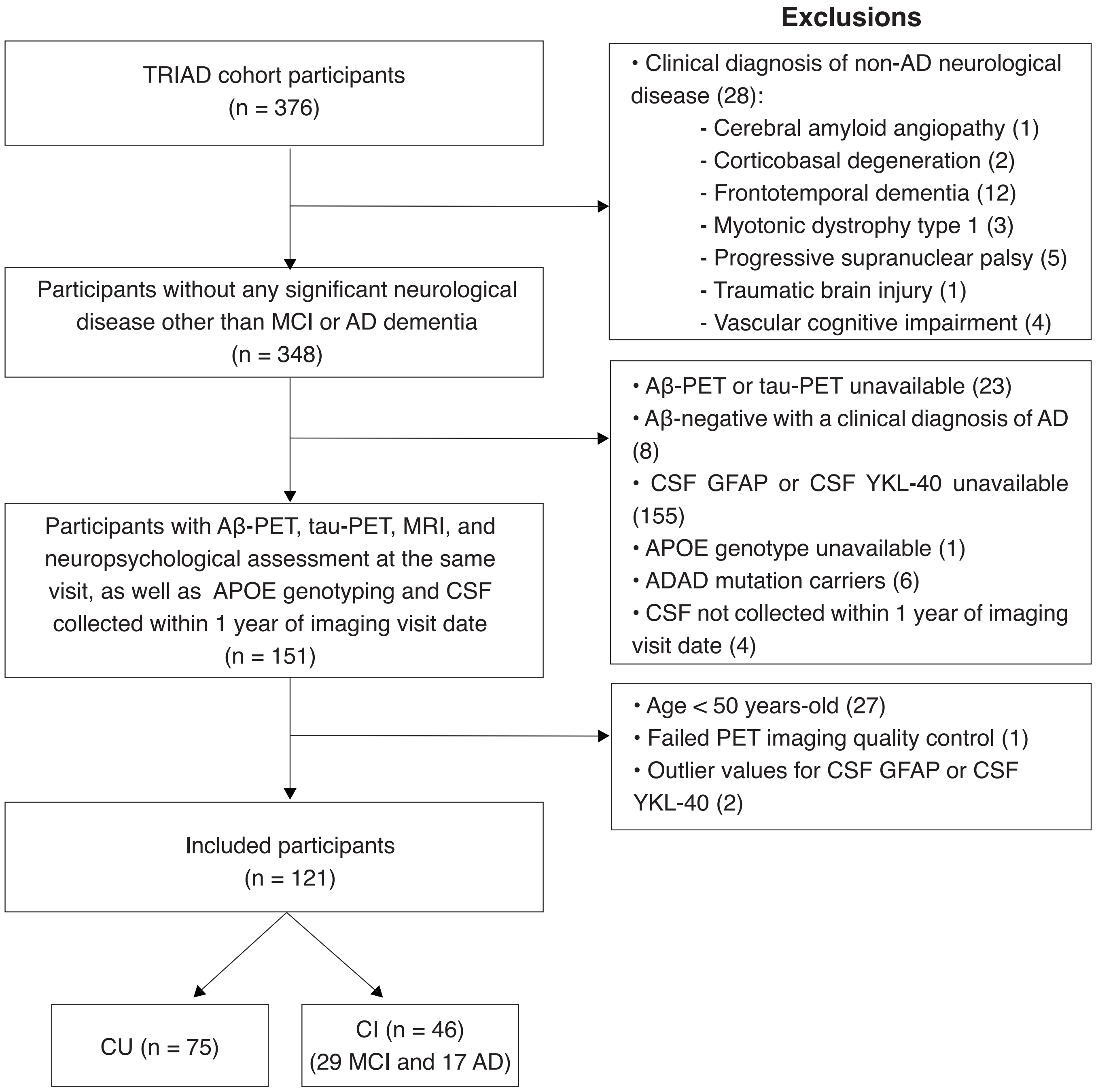


**Supplementary Figure 3. Flowchart of included participants.**

**Supplementary Table 1. Associations of CSF GFAP and YKL-40 with neocortical Aβ-PET and temporal meta-ROI tau-PET.**

| | β (95% confidence interval) | *T*-value | *P*-value |
|---|---|---|---|
| **Model A[a]: neocortical Aβ-PET SUVR ~ CSF GFAP + CSF YKL-40 + temporal meta-ROI tau-PET SUVR + covariates[c]** | | | |
| CSF GFAP | 0.24 (0.08 to 0.40) | 3.03 | 0.003 |
| CSF YKL-40 | -0.14 (-0.30 to 0.03) | -1.66 | 0.100 |
| Temporal meta-ROI tau-PET SUVR | 0.53 (0.33 to 0.72) | 5.38 | < 0.001 |
| **Model B[b]: temporal meta-ROI tau-PET SUVR ~ CSF GFAP + CSF YKL-40 + neocortical Aβ-PET SUVR + covariates[c]** | | | |
| CSF GFAP | -0.08 (-0.22 to 0.06) | -1.17 | 0.244 |
| CSF YKL-40 | 0.24 (0.11 to 0.37) | 3.55 | < 0.001 |
| Neocortical Aβ-PET SUVR | 0.39 (0.24 to 0.53) | 5.38 | < 0.001 |

[a]Adjusted $R^2$: 0.51. [b]Adjusted $R^2$: 0.64. [c]Potential confounders included in the models as covariates are the following: age, sex, cognitive status, and *APOE* ε4 status.

**Supplementary Table 2. Structural equation model coefficients and associated statistics for Figure 2A.**

| | β (95% confidence interval) | *P*-value |
|---|---|---|
| **MMSE** | | |
| Hippocampal volume | 0.28 (0.12 to 0.43) | 0.001 |
| Temporal meta-ROI tau-PET SUVR | -0.48 (-0.68 to -0.28) | < 0.001 |
| Neocortical Aβ-PET SUVR | 0.04 (-0.14 to 0.21) | 0.690 |
| CSF GFAP | -0.02 (-0.16 to 0.12) | 0.762 |
| **Hippocampal volume** | | |
| Temporal meta-ROI tau-PET SUVR | -0.46 (-0.68 to -0.24) | < 0.001 |
| Neocortical Aβ-PET SUVR | -0.03 (-0.23 to 0.17) | 0.783 |
| CSF GFAP | -0.16 (-0.32 to -0.003) | 0.046 |
| **CSF GFAP** | | |
| Temporal meta-ROI tau-PET SUVR | 0.11 (-0.13 to 0.36) | 0.361 |
| Neocortical Aβ-PET SUVR | 0.30 (0.09 to 0.52) | 0.007 |
| **Temporal meta-ROI tau-PET SUVR** | | |
| Neocortical Aβ-PET SUVR | 0.58 (0.45 to 0.70) | < 0.001 |

Model fit indices: CFI = 1.00; RMSEA = 0.00 (90% confidence interval: 0.00-0.00); SRMR = 0.00. All associations were adjusted for age, *APOE* ε4 status, and years of education.

**Supplementary Table 3. Structural equation model coefficients and associated statistics for Figure 2B.**

| | β (95% confidence interval) | *P*-value |
|---|---|---|
| **MMSE** | | |
| Hippocampal volume | 0.30 (0.15 to 0.46) | < 0.001 |
| Temporal meta-ROI tau-PET SUVR | -0.51 (-0.72 to -0.31) | < 0.001 |
| Neocortical Aβ-PET SUVR | 0.03 (-0.14 to 0.20) | 0.706 |
| CSF YKL-40 | 0.09 (-0.05 to 0.23) | 0.198 |
| **Hippocampal volume** | | |
| Temporal meta-ROI tau-PET SUVR | -0.41 (-0.63 to -0.18) | < 0.001 |
| Neocortical Aβ-PET SUVR | -0.08 (-0.27 to 0.11) | 0.418 |
| CSF YKL-40 | -0.17 (-0.33 to -0.01) | 0.041 |
| **CSF YKL-40** | | |
| Temporal meta-ROI tau-PET SUVR | 0.44 (0.20 to 0.67) | < 0.001 |
| Neocortical Aβ-PET SUVR | -0.02 (-0.24 to 0.20) | 0.855 |
| **Temporal meta-ROI tau-PET SUVR** | | |
| Neocortical Aβ-PET SUVR | 0.58 (0.45 to 0.70) | < 0.001 |

Model fit indices: CFI = 1.00; RMSEA = 0.00 (90% confidence interval: 0.00-0.00); SRMR = 0.00. All associations were adjusted for age, *APOE* ε4 status, and years of education.

**Supplementary Table 4. Demographics of the subset of individuals with available CSF inflammation-related proteins.**

| | CU | CI | *P*-value |
|---|---|---|---|
| No. | 36 | 26 | - |
| Age, years | 71.6 (6.1) | 69.6 (7.9) | 0.296 |
| Male, No. (%) | 9 (25.0) | 16 (61.5) | 0.008 |
| Education, years | 14.5 (3.5) | 15.6 (2.8) | 0.174 |
| *APOE* ε4 carriers, No. (%) | 11 (30.6) | 16 (61.5) | 0.030 |
| MMSE score | 29.2 (1.1) | 26.6 (3.9) | 0.002 |
| Neocortical Aβ-PET SUVR | 1.62 (0.5) | 2.30 (0.5) | < 0.001 |
| Temporal meta-ROI tau-PET SUVR | 0.87 (0.1) | 1.54 (0.7) | < 0.001 |
| Hippocampal volume, $cm^3$ | 3.53 (0.3) | 3.24 (0.4) | 0.004 |

Continuous variables are presented as mean (SD).

**Supplementary Table 5. Abbreviations of inflammation-related proteins.**

| Protein symbol | Protein name |
|---|---|
| 4E-BP1 | Eukaryotic translation initiation factor 4E-binding protein 1 |
| ADA | Adenosine Deaminase |
| CCL11 | C-X-C motif chemokine 11 |
| CCL19 | C-C motif chemokine 19 |
| CCL23 | C-C motif chemokine 23 |
| CCL25 | C-C motif chemokine 25 |
| CCL3 | C-C motif chemokine 3 |
| CCL4 | C-C motif chemokine 4 |
| CD244 | Natural killer cell receptor 2B4 |
| CD40 | CD40L receptor |
| CD5 | T-cell surface glycoprotein CD5 |
| CD8A | T-cell surface glycoprotein CD8 alpha chain |
| CDCP1 | CUB domain-containing protein 1 |
| CSF-1 | Macrophage colony-stimulating factor 1 |
| CST5 | Cystatin D |
| CX3CL1 | Fractalkine |
| CXCL1 | C-X-C motif chemokine 1 |
| CXCL10 | C-X-C motif chemokine 10 |
| CXCL11 | C-X-C motif chemokine 11 |
| CXCL5 | C-X-C motif chemokine 5 |
| CXCL6 | C-X-C motif chemokine 6 |
| CXCL9 | C-X-C motif chemokine 9 |
| DNER | Delta and Notch-like epidermal growth factor-related receptor |
| FGF-19 | Fibroblast growth factor 19 |
| FGF-5 | Fibroblast growth factor 5 |
| Flt3L | FMS-related tyrosine kinase 3 ligand |
| HGF | Hepatocyte growth factor |
| IL-10RB | Interleukin-10 receptor subunit beta |

| | |
|---|---|
| IL-12B | Interleukin-12 subunit beta |
| IL-18 | Interleukin-18 |
| IL-18R1 | Interleukin-18 receptor 1 |
| IL-6 | Interleukin-6 |
| IL-7 | Interleukin-7 |
| IL-8 | Interleukin-8 |
| LAP TGF-beta-1 | Latency-associated peptide transforming growth factor beta-1 |
| LIF | Leukemia inhibitory factor |
| LIF-R | Leukemia inhibitory factor receptor |
| MCP-1 | Monocyte chemotactic protein 1 |
| MCP-2 | Monocyte chemotactic protein 2 |
| MCP-4 | Monocyte chemotactic protein 4 |
| MMP-1 | Matrix metalloproteinase-1 |
| MMP-10 | Matrix metalloproteinase-10 |
| OPG | Osteoprotegerin |
| PD-L1 | Programmed cell death 1 ligand 1 |
| SCF | Stem cell factor |
| SIRT2 | SIR2-like protein 2 |
| STAMBP | STAM-binding protein |
| TGF-alpha | Transforming growth factor alpha |
| TNF-beta | TNF-beta |
| TNFRSF9 | Tumor necrosis factor receptor superfamily member 9 |
| TNFSF14 | Tumor necrosis factor ligand superfamily member 14 |
| TRAIL | TNF-related apoptosis-inducing ligand |
| TWEAK | Tumor necrosis factor ligand superfamily, member 12 |
| uPA | Urokinase-type plasminogen activator |
| VEGF-A | Vascular endothelial growth factor A |

### Supplementary References


1 Ashburner, J. A fast diffeomorphic image registration algorithm. *Neuroimage* **38**, 95-113, doi:10.1016/j.neuroimage.2007.07.007 (2007).

2 Ashburner, J. & Friston, K. J. Voxel-based morphometry--the methods. *Neuroimage* **11**, 805-821, doi:10.1006/nimg.2000.0582 (2000).

3 Pascoal, T. A. *et al.* In vivo quantification of neurofibrillary tangles with [(18)F]MK-6240. *Alzheimers Res Ther* **10**, 74, doi:10.1186/s13195-018-0402-y (2018).

4 Cselenyi, Z. *et al.* Clinical validation of 18F-AZD4694, an amyloid-beta-specific PET radioligand. *J Nucl Med* **53**, 415-424, doi:10.2967/jnumed.111.094029 (2012).

5 Pascoal, T. A. *et al.* 18F-MK-6240 PET for early and late detection of neurofibrillary tangles. *Brain* **143**, 2818-2830, doi:10.1093/brain/awaa180 (2020).
